# Excess mortality in the general population versus Veterans Healthcare System during the first year of the COVID-19 pandemic in the United States

**DOI:** 10.1101/2022.09.13.22279868

**Authors:** Daniel M. Weinberger, Liam Rose, Christopher Rentsch, Steven M. Asch, Jesse Columbo, Joseph King, Caroline Korves, Brian P. Lucas, Cynthia Taub, Yinong Young-Xu, Anita Vashi, Louise Davies, Amy C. Justice

## Abstract

**Importance:** The COVID-19 pandemic had a substantial impact on the overall rate of death in the United States during the first year. It is unclear whether access to comprehensive medical care, such as through the VA healthcare system, altered death rates compared to the US population.

**Objective:** Quantify the increase in death rates during the first year of the COVID-19 pandemic in the general US population and among individuals who receive comprehensive medical care through the Department of Veterans Affairs (VA).

**Design:** Analysis of changes in all-cause death rates by quarter, stratified by age, sex race/ethnicity, and region, based on individual-level data. Hierarchical regression models were fit in a Bayesian setting. Standardized rates were used for comparison between populations.

**Setting and participants:** General population of the United States, enrollees in the VA, and active users of VA healthcare.

**Exposure and main outcome:** Changes in rates of death from any cause during the COVID-19 pandemic in 2020 compared to previous years.

**Results:** Sharp increases were apparent across all of the adult age groups (25 years and older) in both the general US population and the VA populations. Across all of 2020, the relative increase in death rates was similar in the general US population (RR: 1.20 (95% CI: 1.17, 1.22)), VA enrollees (RR: 1.20 (95% CI: 1.14, 1.29)), and VA active users (RR: 1.19 (95% CI: 1.14, 1.26)). Because the pre-pandemic standardized mortality rates were higher in the VA populations prior to the pandemic, the absolute rates of excess mortality were higher in the VA populations.

**Conclusions and Relevance:** Despite access to comprehensive medical care, active users of the VA had similar relative mortality increases from all causes compared with the general US population. Factors that influenced baseline rates of death and that mitigated viral transmission in the community are more likely to have influenced the impact of the pandemic.

## INTRODUCTION

During the first year of the COVID-19 pandemic, there was a substantial increase in the rate of death in the United States (US) [1]. This increase, particularly in older adults, was largely a consequence of infection with SARS-CoV-2 [2]. However, the impact of the pandemic varied substantially across subpopulations. Rates of death related to COVID-19 were higher among Black and Hispanic populations compared to White populations, among men compared with women, and among older adults compared with younger individuals [3]. These disparities were influenced by a number of factors including social determinants of health (e.g., employment type, household composition), differing comorbidities, systemic differences in access to healthcare, and biological differences in immune responses by age and sex.

Less understood is the role that the health care system response played in exacerbating or mitigating disparities. A more robust healthcare system might have mitigated some of the worst outcomes of the pandemic in the United States, either by better managing existing chronic health conditions, minimizing disruptions in care, or by an integrated response to the pandemic itself [4]. The US Department of Veterans Affairs (VA) provides comprehensive care nationally for Veterans of the armed forces. Comparing the impacts of the COVID-19 pandemic among VA users to the general US population could shed light on the role that health care played in producing variations in outcomes during the pandemic.

Previous reports show that death rates among those who typically received care in the VA system were less affected than the general US population during the COVID-19 pandemic, despite higher rates of comorbidities [5–7]. However, Veterans enrolled in VA care also differ from the general US population in age, sex, racial/ethnic composition, and geographic distribution, and each of these factors has been associated with mortality rates in general and deaths rates due to COVID-19 specifically [8–10]. Now that individual-level mortality data are available for both populations, it is possible to make appropriate comparisons by accounting for their differing demographic characteristics.

The aim of this analysis was to quantify excess all-cause deaths during the first nine months of the COVID-19 pandemic in Veterans enrolled in the VA system compared with the general US population and to obtain standardized overall estimates of excess deaths, adjusted for age, sex, race/ethnicity, and region.

## METHODS

### Overview

The analyses focused on deaths among adults aged 25 years and older in three different populations: VA enrollees, VA active users, and the general US population. Data were obtained from the National Center for Health Statistics (NCHS) and from the VA for deaths occurring between January 1, 2014 and December 31, 2020. Time series were created by counting the number of deaths by age group, sex, race/ethnicity, year, quarter, and census region (West, South, Midwest, Northeast). A model was fit to quarterly data from 2014-2019 (pre-pandemic) and then extrapolated to 2020. Excess deaths and rate ratios were calculated by comparing the observed number of deaths with the expected number of deaths.’

### Data Sources and Definitions

#### National Center for Health Statistics (NCHS)

Individual-level data representing all deaths in the US were obtained from the NCHS through a data use agreement that allowed for the sharing of geographic location of the deaths [11]. The individual-level data, except for geographic region, are publicly available and can be downloaded from the NCHS website [11]. These vital statistics data have detailed information on the deceased individual including age (years), sex, race/ethnicity, state and county of residence, and the underlying and contributing causes of death (coded using the International Classification of Diseases, 10^th^ revision).

Population size data for the general US population, collected by the US Census Bureau, were obtained from the bridged race files (Vintage 2020) produced by the NCHS. These data are available at the annual scale (July 1 estimates). To estimate population size for each stratum by quarter, we used linear interpolation.

#### Department of Veterans Affairs (VA)

The VA enrollee population includes approximately 10.9 million individuals enrolled in VA health care: 9.2 million Veterans and1.7 million who are family members of disabled Veterans [12]. The population of the VA system is commonly defined as either all enrollees or those who actively use the VA system. The majority of enrollees have another form of health insurance, with Medicare being the most common among those aged 65 years or older [13]. The baseline comorbidities and rates of deaths vary between these populations, and as such we include two definitions in our analysis (i.e., “VA enrollees”, “VA active users”).

VA enrollees were defined as any individuals eligible for care, either paid for or provided by the VA. VA active users were individuals with at least one diagnosis in their VA electronic health record in the two years prior to each time point, indicating at least one clinical encounter within the VA [14]. The VA active user cohort is dynamic, with the population determined by activity in the previous two year from each time point.

Population size data for the VA populations were obtained by querying the Assistant Deputy Under Secretary for Health (ADUSH) Enrollment Files for each fiscal year.

#### Definitions

These analyses focused on deaths among adults aged 25 years and older. The three populations are nested within each other (VA active users are also VA enrollees; VA enrollees are also part of the general US population). The analyses focused on deaths among adults aged 25 years and older because there are very few Veterans under the age of 25 (1.3% of Veterans).

For each population, we used the same definitions for creating subgroups:

- Age group: (25-44, 45-64, 65-74, 75-84, 85+ years)
- Sex (male/female)
- Race/Ethnicity: non-Hispanic White; Non-Hispanic Black or African American; Hispanic; American Indian/Native Alaskan; Asian, Native Hawaiian and Other Pacific Islander. These groups were chosen to align with population size data available from the National Center for Health Statistics (NCHS) [16].
- Quarter (Jan-Mar, Apr-Jun, Jul-Sep, Oct-Dec)
- Census region (Northeast, South, West, Midwest) [15],

### Statistical analysis

#### Model to generate a baseline

The goal for the model was to generate an expected number of deaths by month and subgroup based on pre-pandemic quarterly data from the preceding six pre-pandemic years (2014-2019) and then extrapolate to each quarter in 2020. Because the pandemic began in earnest in the US in March 2020, the first quarter of 2020 includes both pandemic and pre-pandemic months and is therefore not used for model fitting. Excess deaths and rate ratios are calculated by comparing the observed number of deaths with the expected number of deaths based on the model. Observed and modeled values were combined over subgroups to obtain summary estimates. We used a hierarchical regression model fit in a Bayesian setting using the INLA packaged in R *(Supplementary Methods*).

#### Standardized death rates

For comparisons between the different populations, we use death rates standardized by age, sex, race/ethnicity, and region, using direct standardization. The population of VA enrollees who have non-missing data on race/ethnicity was used as the standard population. Both the observed and expected deaths rates were standardized, and the ratio of these standardized values provides a mortality rate ratio adjusted for population structure.

#### Individual-level analysis

We leveraged the full patient-level data from VA active users to compare estimates of excess mortality using the Poisson model described above, which are fit to aggregate data, with a patient-level Cox proportional hazards model, fit to individual-level data (Supplementary Methods).

#### Availability of code and data

Code used for all analyses can be found at https://github.com/VA-CareDisruptions/VA_CDC_death_comparison. All analyses were completed in R [17]. Data on US mortality, with the exception of state/region, can be obtained from https://www.cdc.gov/nchs/nvss/mortality_public_use_data.htm. For additional variables, including geography, a data use agreement with NCHS is required. VA data and the analytic data sets used for this study can be made available to researchers with a VA IRB approved study protocol and data use agreement. Information is available at https://www.virec.research.va.gov or contact the VA Information Resource Center at.

## RESULTS

### Demographics and baseline death rates

The demographics of the VA population differed from those of the general US population in several important ways. The VA population was predominantly male, older than the general US population, and had a larger proportion who were Black or Hispanic (**Table 1**). Expected death rates were generally higher in the VA enrollees than the general US population (2520 vs 930 deaths per 100,000) and higher still in active users of VA healthcare (2910 deaths per 100,000) (**Table 2**). Even after standardizing the death rates by age, sex, race/ethnicity, and region, the expected death rate among the VA active users was ∼25% higher than in the general US population (2160 deaths/100,000 in the US population vs 2730 among VA active users (**Table 2**).

**Table 1:**
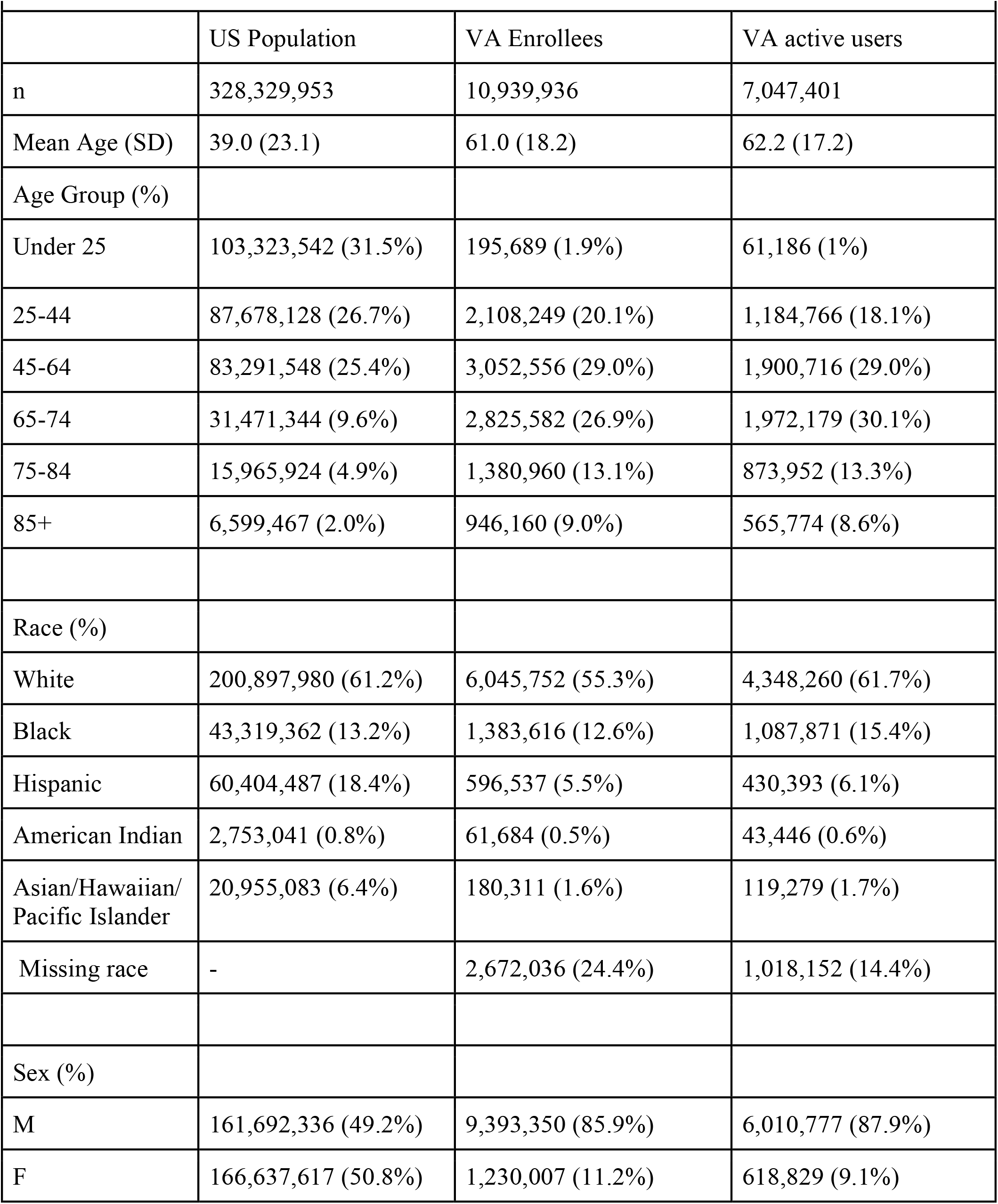
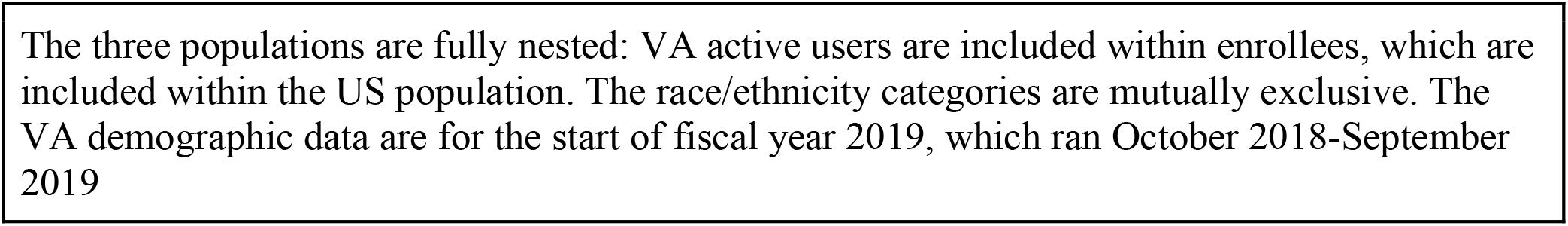
Characteristics of the Populations in 2019

**Table 2.**

| Table 2. |  |  |  |
| --- | --- | --- | --- |
|  | United States | VA enrollees | VA active users |
| All-cause Deaths/100,000* |  |  |  |
| Expected 2020 Q2-4 | 930 (910, 960) | 2520 (2360, 2670) | 2910 (2770, 3050) |
| Observed 2020 Q2-4 | 1130 | 3022 | 3441 |
| Excess incidence | 200 (170, 220) | 500 (360, 660) | 530 (95%CI:390, 680) |
| Rate ratio | 1.22 (1.18, 1.25) | 1.20 (1.13, 1.28) | 1.18 (1.12, 1.24) |
| Reported COVID deaths/100,000 | 170 | - | - |
| Deaths/100,000* people (standardized to VA enrollee population) |  |  |  |
| Expected 2020 Q2-4 | 2160 (2130, 2200) | 2360 (2210, 2500) | 2730 (2590, 2860) |
| Observed 2020 Q2-4 | 2588 | 2842 | 3256 |
| Excess incidence | 430 (390, 460) | 480 (350, 630) | 530 (400, 670) |
| Rate ratio | 1.20 (1.17, 1.22) | 1.20 (1.14, 1.29) | 1.19 (1.14, 1.26) |
| * All mortality rates calculated as deaths per 100,000 people age 25 and above during 2020 Q2-Q4. Expected incidence and excess incidence are rounded to the nearest 10. |  |  |  |

### Relative and absolute increases in rates of death in 2020

There were sharp differences between rates of observed and expected deaths due to any cause starting in the 2^nd^ quarter of 2020 (April onwards). These increases were apparent across all of the adult age groups (25 years and older) in both the general US population and the VA populations (**Figure 1)**. There were subtle differences in the magnitude of the relative increases during the first wave of the pandemic (2020 Q2), with a larger increase in the general US population than either of the VA populations. (**Figure 2A**) However, these differences were largely due to differences in the geographic and demographic makeup of the populations, which were resolved by standardization (**Figure 2B, Figure S1**). Across all of 2020, the relative increase in death rates during 2020 was similar in the general US population (RR: 1.20 (95%CI: 1.17, 1.22)), VA enrollees (RR: 1.20 (95%CI: 1.14, 1.29)), and VA active users (RR: 1.19 (95% CI: 1.14, 1.26)). Because the pre-pandemic standardized mortality rates were higher in the VA populations prior to the pandemic, the absolute rates of excess mortality were higher in the VA population despite the similar relative increases in all three populations (**Figures 2 and 3, Table 2**).

**Figure 1.**
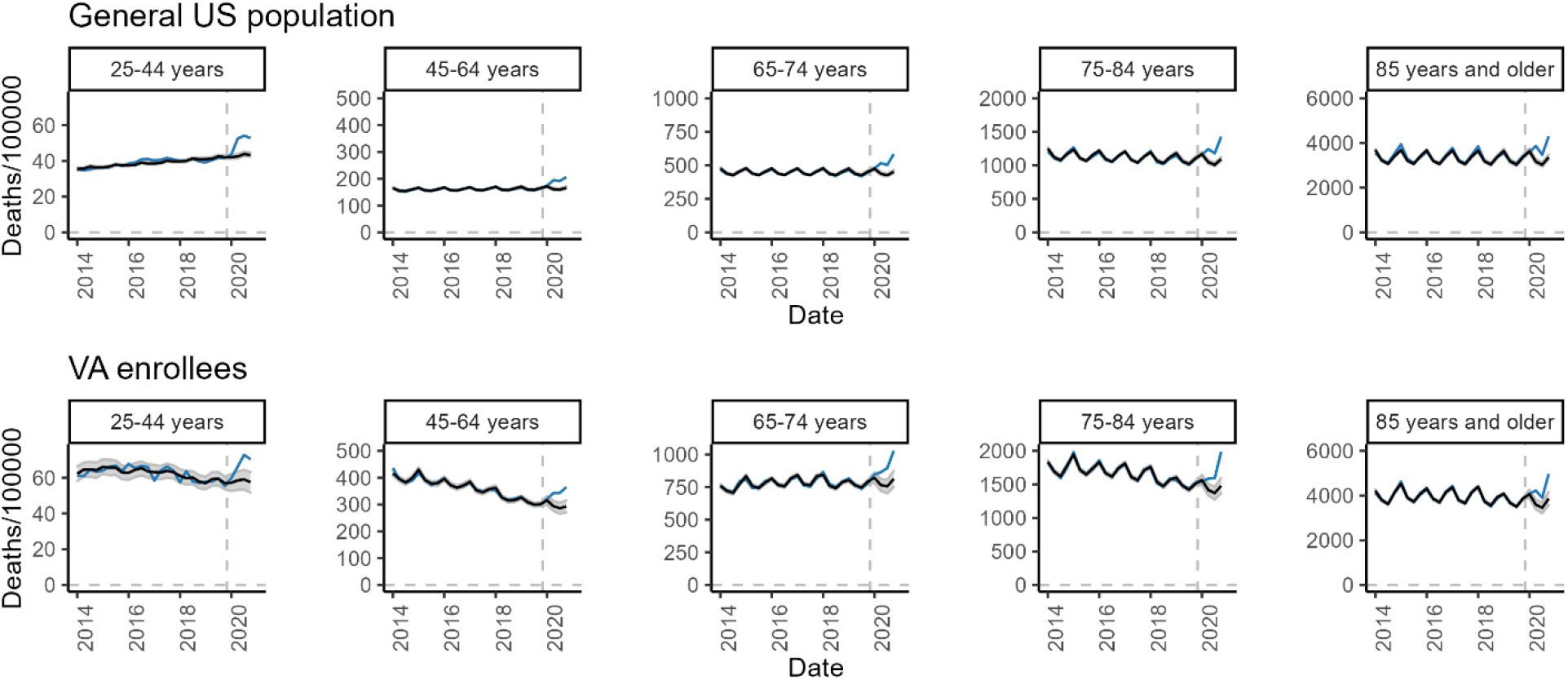
Observed and expected rates over time by age group among the general US population and among enrollees of the VA. The Black line with the shaded area indicates the fitted values (up through 2019) and predicted values (for 2020), while the colored lines show the observed incidence by quarter.

**Fig 2:**
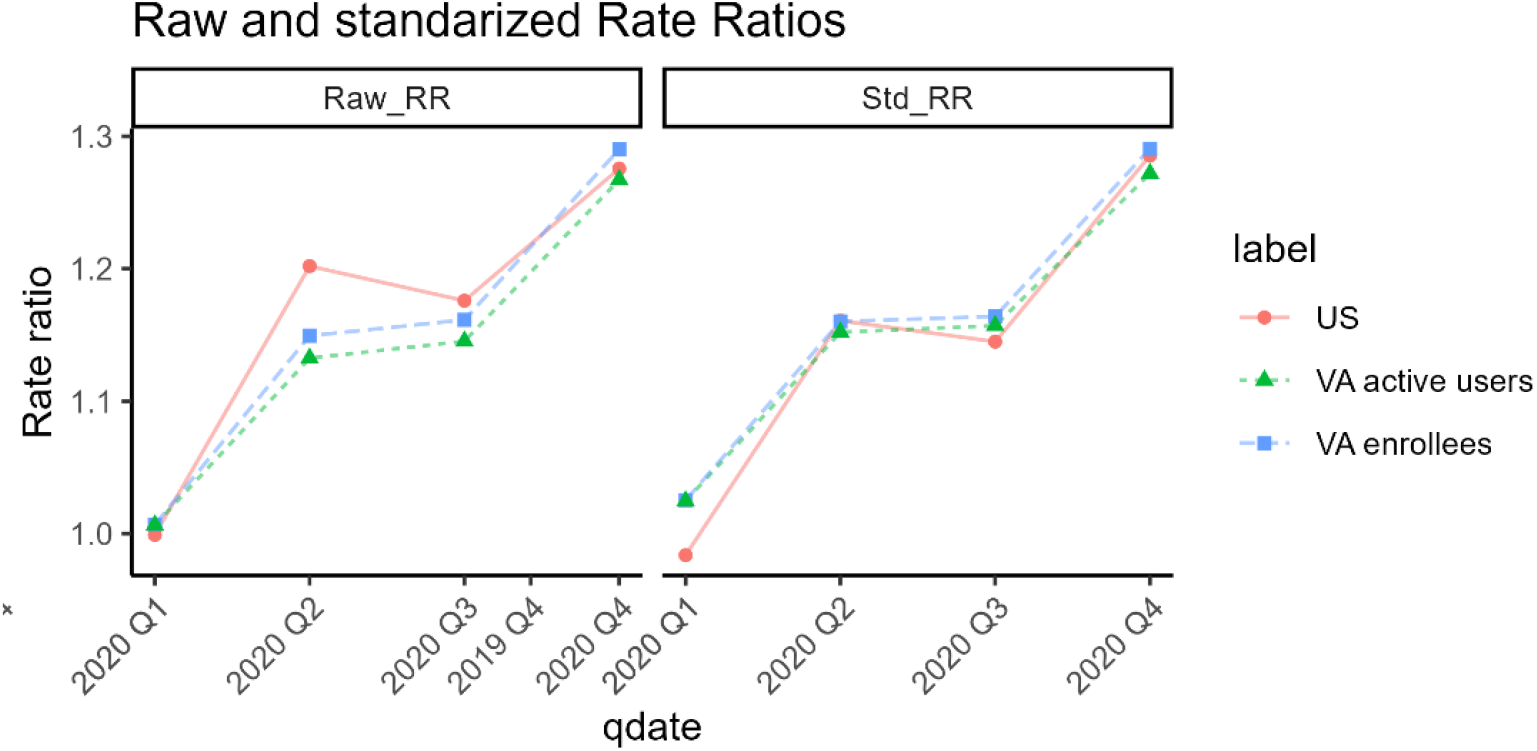
Trajectory of the rate ratio for the US, VA enrollees, and VA active users, calculated using (A) the raw mortality rates or (B) the mortality rates standardized by age, sex, race/ethnicity, and region. The closer alignment for the standardized plot on the right suggests that any differences on the left panel are related to demographic differences.

**Fig 3.**
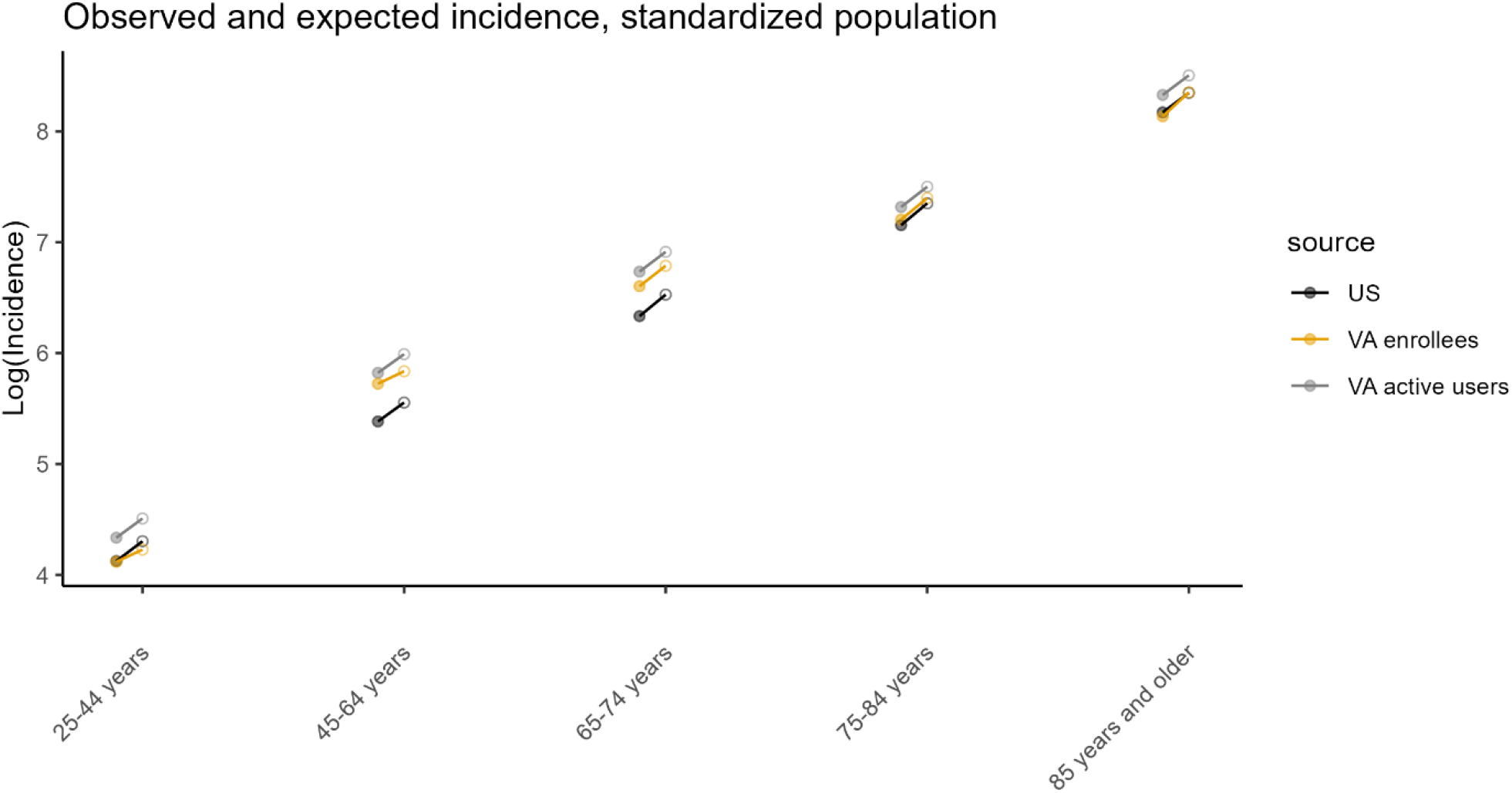
Observed mortality rates (open symbols) and expected mortality rates (closed symbols) in the US population, VA enrollees, and VA active users, stratified by age. The slope of the line indicates the relative increase during the pandemic (the rate ratio). Mortality rates are standardized based on the sex, race/ethnicity, region and distribution of the 65-79 year old VA enrollee population

### Comparison of excess death rates by age, sex, and race/ethnicity

While women comprised a small fraction of the VA population, the relative increase in rates of death during 2020 was more notable for women compared to men among VA enrollees, particularly among younger women (**Figure S2**). These estimates are based on small numbers and should be interpreted with caution. The relative increase in deaths was smaller in the White population than in other racial/ethnic groups, and this was consistent between the general US population and the VA populations (**Figure S3)**.

### Rates of excess deaths vs recorded COVID deaths

Overall, in 2020 there were 200 excess deaths/100,000 people in the US population (95%CI: 170, 220). During the same period, there were 170 deaths/100,000 people that were recorded as having COVID-19 as the underlying or contributing cause (**Table 2**). This suggests that ∼85% of the increase in the death rate was directly attributable to COVID-19 (170 COVID-19 deaths/200 total excess deaths). Access to specific causes of death were not available for the VA population unless the individual received a test for SARS-CoV-2 within the VA system.

### Individual-level analysis

Estimates of the relative increase in death rates during 2020 among VA active users was similar between the Poisson model (RR 1.18, 95% CI 1.15-1.22) and the patient-level Cox model (RR 1.17, 95% CI 1.15-1.20). There was also no meaningful difference in the estimates by quarter (**Table S1**).

## DISCUSSION

In this study, we find that after adjusting for differences in age, sex, race/ethnicity, and region, the relative increase in rates of death during the first nine months of the COVID-19 pandemic were similar between the general US population and enrollees and users of the VA healthcare system. The absolute pre-pandemic death rate was higher among the VA populations, and this translated to higher absolute excess death rates among Veterans, despite the similar relative increases.

A key question is whether having access to a unified healthcare system through the VA affected the risk of death during the first phase of the pandemic. Our analysis demonstrates that the COVID-19 pandemic effectively acted as a multiplier on the baseline death rates. Therefore, if individuals receiving care in the VA system had a lower risk of death than if they had not had access to care, then the pre-pandemic death rate might have been even higher, and the rates of excess deaths might have been higher as well. The higher baseline rates of death in the VA population were expected because those in the VA system, particularly the active users, are enriched for those seeking regular medical care, who might be more likely to have comorbidities, which is consistent with the results of prior investigators [18]. To evaluate whether the VA reduces baseline rates of death, it would be necessary to evaluate death rates among individuals outside of the VA system with similar comorbid conditions.

There are also reasons to expect that access to unified healthcare might not be impactful. Early in the pandemic, vaccines and effective treatments were not yet available, so access to healthcare for COVID-19 might not have been a major driver of outcomes. In the first year, most of the excess deaths were likely due to COVID; moving forward into subsequent years of the pandemic, when there were more treatment and prevention strategies, having access to quality care could have a larger effect. Additionally, while processes of care tend to be better in the VA system [19,20], access to care, wait times for procedures, and outcomes are similar in the VA and in non-VA healthcare systems [21,22].

The increase in death rates during the pandemic differed by racial/ethnic group, and this was consistent between the three populations. Previous work demonstrated that once hospitalized, Black and White Veterans had similar rates of death due to COVID-19 [23]. This suggests differences in the impact of the COVID-19 pandemic are due to other factors, primarily the rate of infection in the different groups. The risk of infection varied by race/ethnicity and region [24], and larger household size was also associated with greater infection risk [25].

While the numbers were small, the data suggested that the increase in deaths during the pandemic was more acute for women in the VA system, and particular young women. Women make up a small percentage of the overall VA population and are more predominantly people of color. They could differ from women in the general population in terms of their occupations and exposures to the virus. The female population was too small to evaluate interactions between race/ethnicity and gender.

Measuring excess mortality has advantages and disadvantages as an analytical approach for understanding the impact of the pandemic. By evaluating the overall increase in rates of death, regardless of cause, issues related to variations in viral testing and difference in cause-of-death coding practices are mitigated. The downside is that there is not a perfect correlation between excess deaths and deaths caused by COVID-19. The magnitude of the excess deaths could result directly from deaths caused by the virus, or they could indirectly result from a number of factors including avoidance of emergency care services, disruptions to routine or emergency care, changes in rates of other infectious diseases (e.g., the disappearance of influenza in 2020), and changes in rates of violent crime and overdoses that might have been related to pandemic disruptions [2]. The magnitude of the increase in death rates in the US was similar to the magnitude of recorded COVID-19 deaths (170 recorded COVID-19 deaths per 100,000 vs 200 overall excess deaths), suggesting that much of the increase is directly related to the virus. Modeling studies that make use of the time series of deaths suggest that most of the increase in death rates in older adults was directly related to the virus, based on the timing and trajectory of the increases in all-cause deaths [2]. It is likely that many of the deaths in younger adult age groups were not directly related to the virus, so other indirect factors, including worsening mental health and increases in substance abuse, are likely to have a larger relative impact in younger age groups [2]. Analysis of the data using Poisson models and individual-level hazard models yielded similar estimates given adjustment for the same set of baseline covariates, as would be expected. Future work could focus on further developing the individual-level models to account for additional indicators of underlying health status. Further, longer term mortality consequences of delays in treatment for older individuals with comorbid diseases may take longer to be observed.

These analyses have some important limitations. Race/ethnicity data were missing for 24% of VA enrollees and 7-14% of active users. The data for these individuals is effectively dropped when calculating the standardized rates. If the race/ethnicity data are missing at random, this would not introduce bias into the estimates. However, individuals with missing race/ethnicity information tend to be less dependent on VA care and are more likely to be non-White [26]. Therefore, we likely under-count the impact of the pandemic on non-White populations. While we are able to standardize comparisons by age, sex, race/ethnicity, and region, we did not have individual-level data on comorbidities in the general US population. Therefore, differences in the prevalence of comorbidities between Veterans and general US populations that do not correlate with the adjusted factors could confound the comparisons [27]. Estimates of excess deaths depend on the statistical modeling, including trend and seasonal components, being correctly specified. We used a hierarchical model, which generally will generate a more stable model for sparse groups, but these types of models can also introduce bias for individual groups. This could lead to over- or under-estimation of the trend for some groups, causing inaccurate estimates of excess deaths for subgroups.

In conclusion, using detailed mortality records, we find that the relative impact of the COVID-19 pandemic on all-cause mortality was similar in the general US population and among users of the VA healthcare system. This suggests that access to comprehensive healthcare alone was not sufficient to mitigate the worst impacts of the virus during the first 10 months of the COVID-19 pandemic.

**Table S1.** Estimated rate ratio from individual-level and ecological models fit to data on VA active users

| Model type | IRR/HR Overall |  | IRR/HR Q1 | IRR/HR Q2 | IRR/HR Q3 | IRR/HR Q4 |
| --- | --- | --- | --- | --- | --- | --- |
| Patient (Cox) | 1.17<br>(1.15, 1.20) |  | 1.02<br>(0.99, 1.05) | 1.15<br>(1.12, 1.18) | 1.19<br>(1.16, 1.22) | 1.35<br>(1.32, 1.39) |
| Ecologic (Poisson) | 1.18<br>(1.15, 1.22) |  | 1.00<br>(0.96, 1.04) | 1.10<br>(1.06, 1.15) | 1.15<br>(1.10, 1.20) | 1.29<br>(1.24, 1.35) |

**Fig S1:**
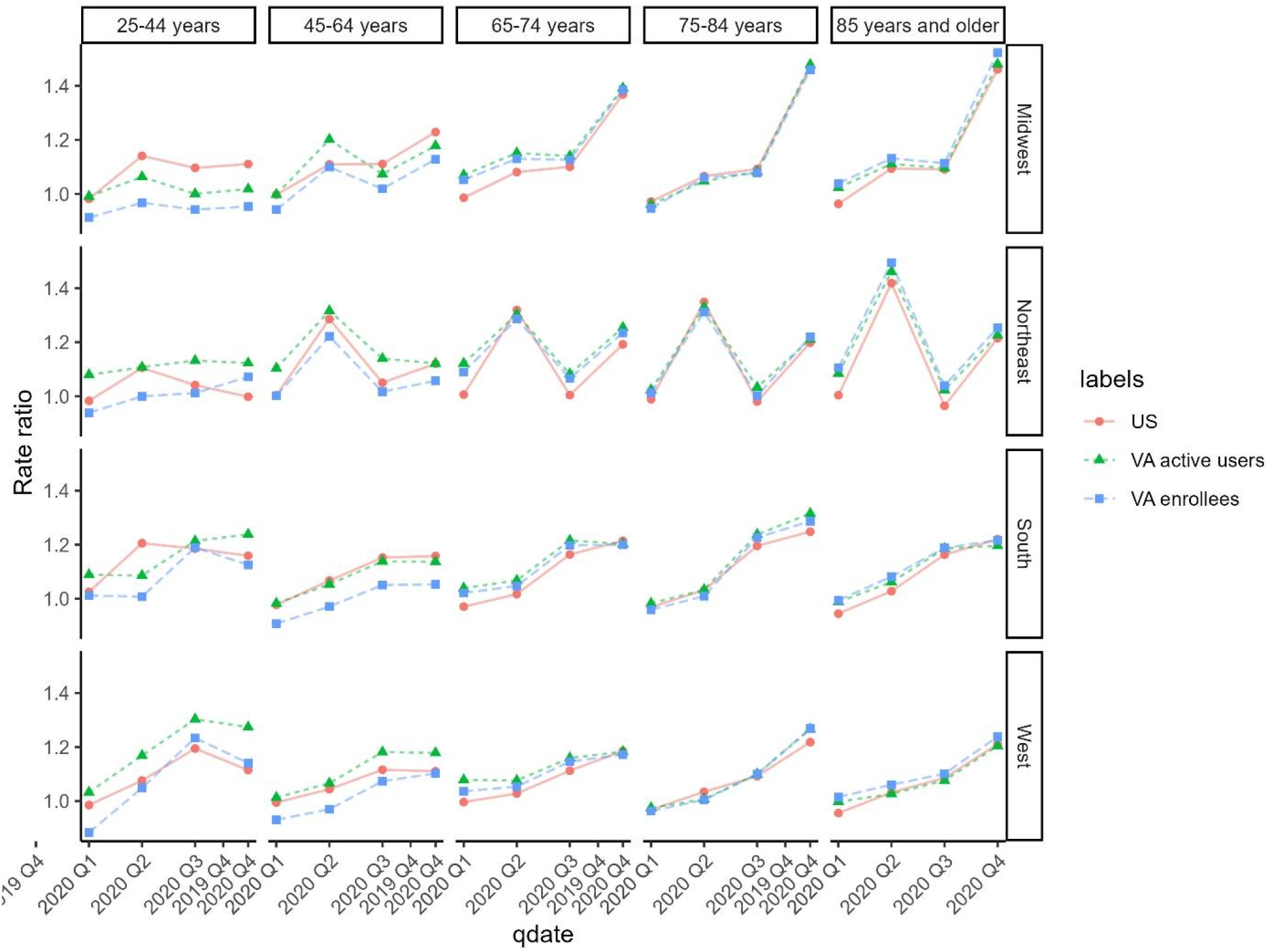
Standardized rate ratio by quarter in 2020 (observed/expected), stratified by age and region, among White men.

**Fig S2.**
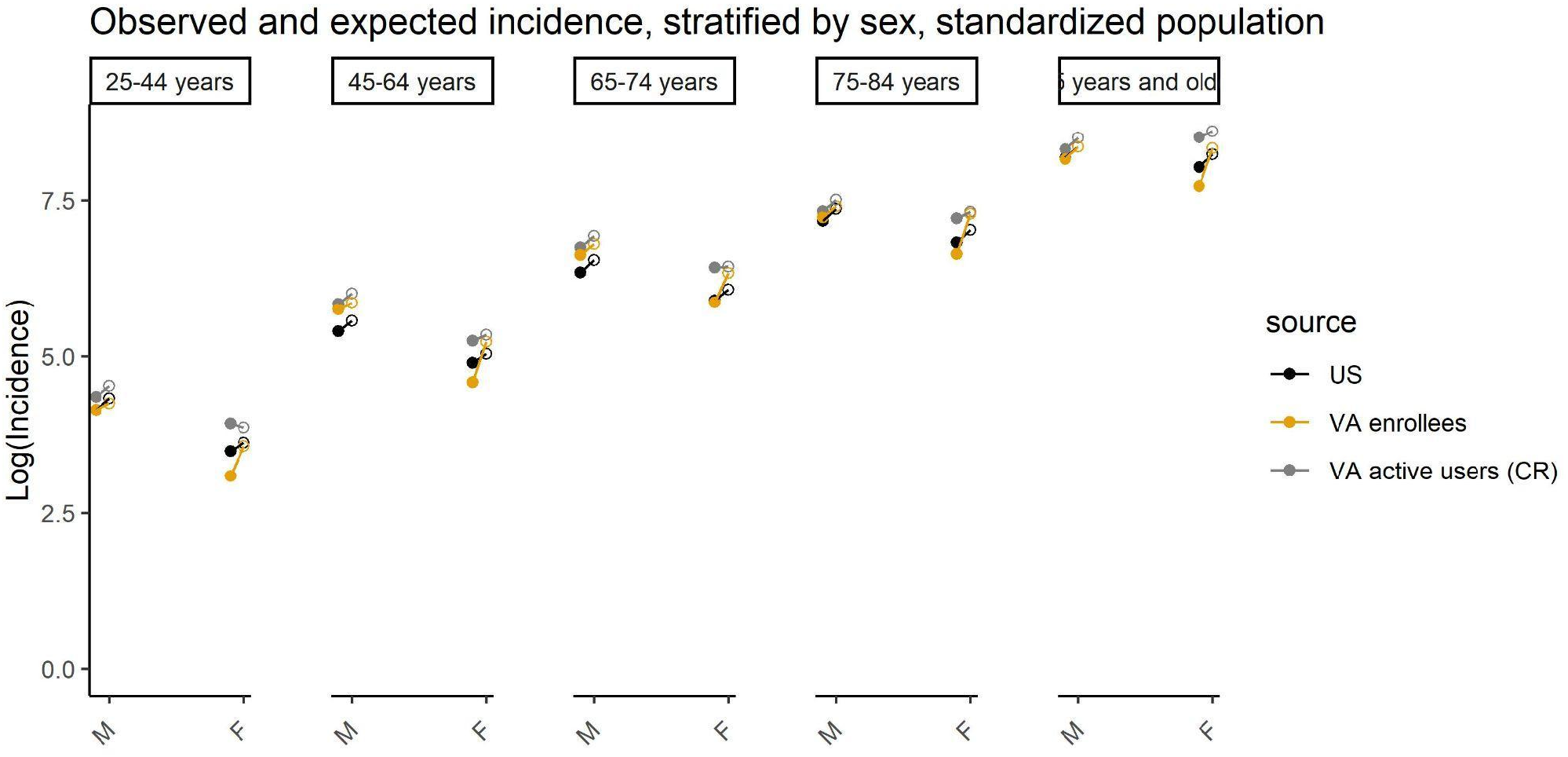
Observed mortality rates (open symbols) and expected mortality rates (closed symbols) in the US population, VA enrollees, and VA active users, stratified by age and sex. Slope of the line indicates the relative increase during the pandemic. Mortality rates are standardized based on the region and race/ethnicity distribution of the 65-74 year old VA enrollee population

**Fig S3.**
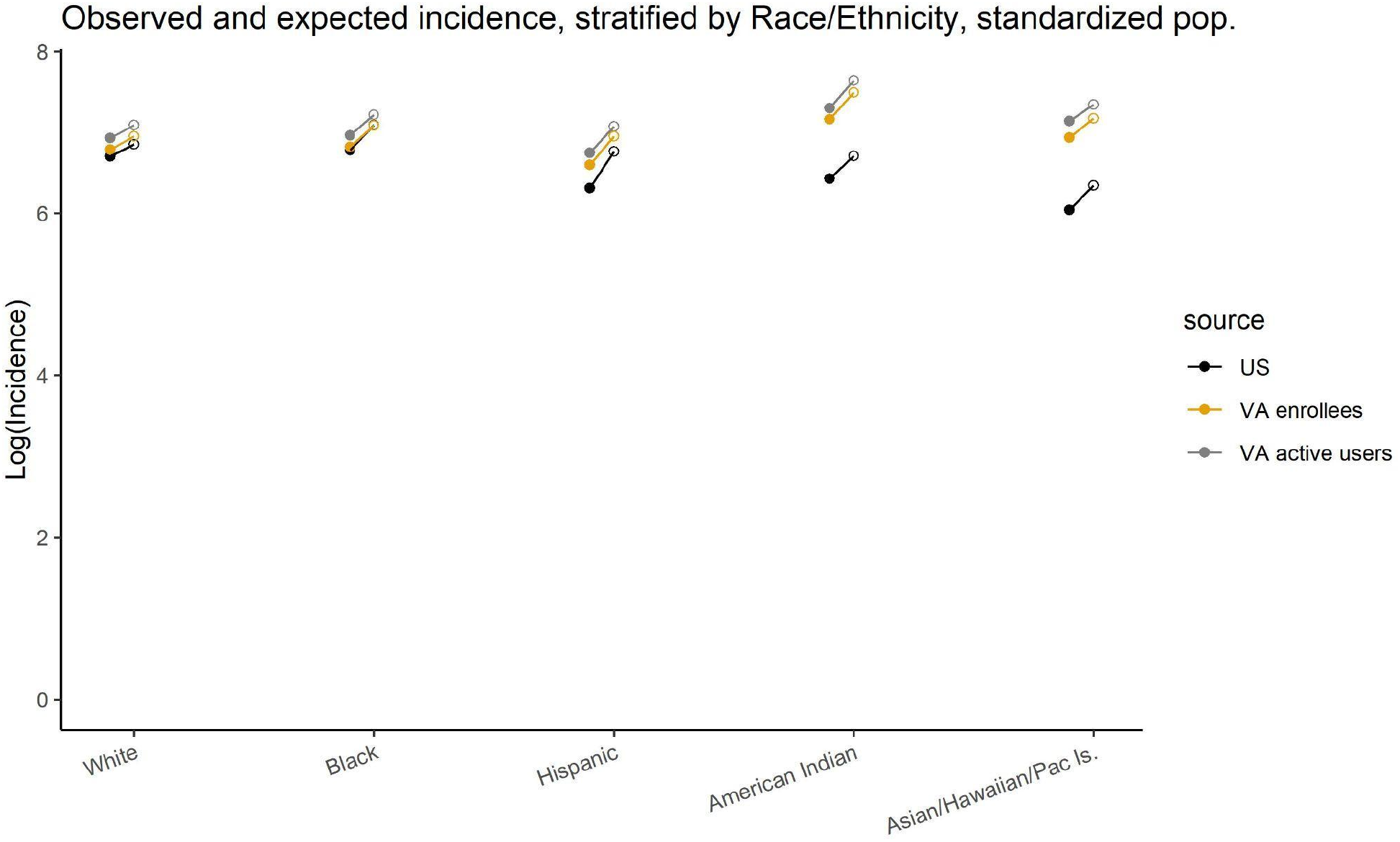
Observed mortality mortality rates (open symbols) and expected mortality rates (closed symbols) in the US population, VA enrollees, and VA active users, stratified by race/ethnicity. Slope of the line indicates the relative increase during the pandemic. Mortality rates are standardized by age, sex, region, and race/ethnicity to the VA enrollee population.

## Data Availability

Code used for all analyses can be found at https://github.com/VA-CareDisruptions/VA_CDC_death_comparison. Data on US mortality, with the exception of state/region, can be obtained from https://www.cdc.gov/nchs/nvss/mortality_public_use_data.htm. For additional variables, including geography, a data use agreement with NCHS is required. VA data and the analytic data sets used for this study can be made available to researchers with a VA IRB approved study protocol and data use agreement. Information is available at https://www.virec.research.va.gov or contact the VA Information Resource Center at.

## Acknowledgement

This material is based upon work supported by the Department of Veterans Affairs, Veterans Health Administration, Office of Research and Development, Health Services Research and Development.

## SUPPLEMENTARY TEXT

### Supplementary Methods

#### Model details

Data in any given subgroup are sparse, making it difficult to fit separate models for each subgroup (as was done in previous work [28]). Instead, we used a hierarchical Bayesian analysis approach. With this approach, we fit a model to disaggregated data, but parameters were estimated hierarchically so information was shared between strata. The model adjusted for seasonality using categorical variables for quarter, adjusted for linear time trends, and uses a population offset to adjust for the size of the at-risk population. An AR(1) random intercept was included to capture unexplained variability in the data that was common across the different strata. The model for the number of deaths in age group *i*, race/ethnicity group *j*, sex *k*, region *r*, and time *t* was:

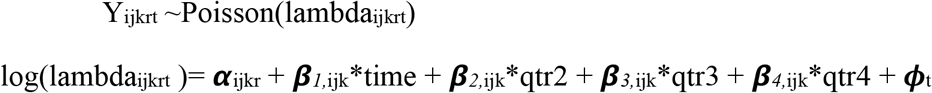

The intercept for each subgroup is given as

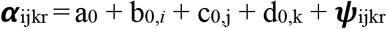

The trend and effect of seasonality for each subgroup is given as

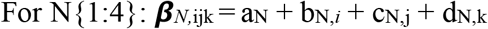

*ϕ*_t_ is an AR(1) random intercept, which captures unexplained temporal variation that is shared across the different subgroups. a_x_, b_x_, c_x_, d_x_ represent the contributions to the intercept, slope and quarterly effects by age group (b), race/ethnicity group (c), and sex (d). A separate intercept (*α*_ijkr_) was estimated for each region, but the time trends and seasonal effects (*β*_*N*,ijk_) were shared across regions. These models were fit using the INLA package in R, which uses approximate Bayesian inference. Samples from the joint posterior distribution were obtained using the inla.posterior.sample() function. Samples were combined across subgroups for calculating summary measures and uncertainty intervals.

The a_N_ are assigned weakly informative priors (N(0,1)); the b_N_, c_N_, and d_N_ have shrinkage priors (N(0, tau_m_)) and use the ‘Z-model’ formulation for random effects in INLA. Models were fit separately to the time series for the different populations (e.g., US population, VA active users).

#### Individual-level analysis

We leveraged the full patient-level data from VA active users to compare estimates of excess mortality using the Poisson model described above, which are fit to aggregate data, with a patient-level Cox proportional hazards model, fit to individual-level data (Supplementary Methods). To reduce computational burden, we limited the pre-pandemic time period in this post-hoc analysis to 2 years (i.e., 2018-2019) for both the individual-level and aggregate models, and a linear trend was not included due to the short baseline period. The Cox model was specified using the same variables as the Poisson models and used current age as the time-scale.

